# Secondhand Cannabis Smoke Exposure: Prevalence, Personal Use, and Neurocognitive Trajectories Over Time in Adolescents in the United States

**DOI:** 10.64898/2026.08.31.26361835

**Authors:** Jenicca Bastien, Kimberly Garcia, Alexander L. Wallace, Ryan M. Sullivan, Eunha Hoh, Natasha E. Wade

## Abstract

**Background:** As cannabis policy changes in the United States, secondhand cannabis smoke (SCS) is increasingly common, including within families. However, prevalence of exposure and clinical correlates over time in adolescents are not fully understood.

**Objectives:** (1) To estimate the prevalence of SCS and personal cannabis use in US-based teens exposed to SCS, and (2) examine the cognitive trajectories of adolescents exposed to SCS compared to non-exposed peers.

**Methods:** Data from the Adolescent Brain Cognitive Development (ABCD) Study was used. Participants (n=11,316 of full cohort with follow-up data; n=776 with self-reported family SCS exposure) attended yearly visits from ages 11-17, completing substance use interviews, toxicological testing, and the NIH Toolbox Cognitive battery. Youth with SCS but no personal cannabis use (n=419; 47% female) were matched on prenatal substance exposure, family substance use history, and sociodemographics to non-SCS exposed and non-cannabis-using youth with a 1:2 ratio (Controls n=838). Linear mixed-effects models assessed cognitive performance by SCS*age interactions, accounting for random effects of subject and family. Covariates included sex and alcohol, nicotine, and other substance use. Secondary models analyzed performance by cumulative waves of reported SCS exposure interacting with age.

**Results:** Of the full cohort, 6.9% (n=776) reported exposure to SCS. Of these individuals, 46% endorsed lifetime personal cannabis use by age 17, relative to 20% of non-SCS exposed youth (OR=3.83[95%CI:3.29,4.44]). Within matched participants, SCS*age demonstrated a significant interaction on attention and inhibitory control (β=-0.32, *p*=.028), with SCS demonstrating reduced improvement over time. More waves of exposure were also associated with worse performance over time (β=-0.39, *p*=.057).

**Discussion:** Almost half of those who had been exposed to SCS endorsed personal cannabis use. Cognitive findings were domain specific, similar to findings in secondhand tobacco: SCS exposed youth showed restricted improvement in attention and inhibitory control by age 17. Public health and policymakers should make efforts to curb youth SCS exposure, given the potential for risk which has not been fully explored to date.

## Introduction

Cannabis use is increasingly popular, with 24 states legalizing recreational use in the United States by 2026. The most common route of cannabis use in U.S. adults is smoking, followed by eating and vaping ^1,2^. Of those who smoke cannabis over 60% reported indoor use in the 2020 Global Drug Survey ^3^. Concerningly, in-home cannabis smoke is now more prevalent than in-home tobacco smoke ^4^, with significantly increasing rates among parents over the past 20 years ^5,6^. In fact, over a quarter of those who smoke at home do not restrict their use when minors are present ^7^. The impacts of exposure to secondhand cannabis smoke (SCS) are limitedly understood, thus more research is warranted to understand possible cognitive relationships.

Research suggests that familial substance use is a predictor of personal substance use ^8^. Parents who used cannabis frequently during their own adolescence, were more likely to raise children with earlier onset of cannabis use ^9^. Young adults with parents who used cannabis (at any point in their life) were more likely to report cannabis use themselves, in addition to earlier age of initiation and more frequent use ^10,11^. Furthermore, youth involvement in familial substance use (e.g., lighting a cigarette for a parent) is associated with personal use of those substances concurrently and within one year ^12^. Adolescence is a common period of substance use initiation ^13^, though the relationship between SCS exposure and rates of personal use are not fully known.

Although the physiological and cognitive effects of personal cannabis use are widely studied ^14–16^, there is limited research on exposure to SCS. In a pre-clinical study, Moir et al. ^17^ found known carcinogens and chemicals that contribute to respiratory diseases in SCS. Additionally, SCS has higher particulate matter than secondhand tobacco smoke (STS) ^18^. Similar findings have been shown in other studies when looking at cannabis smoke from a marijuana cigarette and a bong ^19,20^. These environmental contaminants may contribute to brain-behavior relationships, which warrants further follow-up research in youth samples to consider potential clinical correlates of exposure.

While STS contributes to known physical and cognitive impairments in youth ^21,22^, it is unclear whether SCS influences similar trajectories. In one of the few studies on SCS, young children (ages 1-5) exposed to SCS showed more aggressive and defiant behavior, as well as poorer cognitive flexibility and language reception when parents reported in-home cannabis exposure, even after accounting for prenatal exposure ^23^. Additionally, there are higher rates of asthma in teenagers with SCS ^24^, and respiratory symptoms in adults after exposure to SCS ^25^. These physical effects may have downstream brain and cognition impacts. To that end, our group previously found that SCS-exposed participants had a negative marginal relationship between visuospatial performance but performed better on an oral reading task in preadolescents ages 11- 12 ^22^. However, our previous study had a relatively small sample size, with only 88 participants examined cross-sectionally, necessitating longitudinal analysis and a larger sample.

In this study we first sought to estimate the prevalence of SCS and personal cannabis use in teens exposed to SCS in a longitudinal cohort, hypothesizing that those with SCS exposure would demonstrate higher rates of personal cannabis use. Our second aim is to investigate the relationship between SCS and cognition over time in adolescents (ages 10-17). We consider in-home SCS exposure, comparing any reported exposure over 5 waves, to well matched participants without SCS exposure, expecting those with SCS exposure to demonstrate reduced improvement in cognition over time. We also examine whether the number of waves of exposure (0-5) dimensionally predicts cognition, expecting more waves of exposure to be associated with worse performance. Across models, we robustly control for potential confounds through matching on sociodemographics, prenatal exposure, parent history of substance use problems, and STS exposure, while excluding for personal cannabis use and accounting for time-varying other substance use.

## Methods

Participants (n=11,316) were 10.62-17.8 across waves of included data (Years 2-6) from the annual ABCD Study data release 7.0 <u>(DOI 10.1.66.93/8f3w-5260)</u>. Prevalence rates were estimated from the full cohort, with cognitive correlate hypothesis testing in a subset (n=776 with secondhand cannabis smoke (SCS) exposure; n=419 with SCS without personal cannabis use and included in cognitive modeling). ABCD began recruitment of children ages 9-10 in 2016 with the goal of recruiting up to 11,500 children across 21 sites ^26,27^. The study conducts annual assessments including cognitive assessments, and a range of questionnaires including surveys on substance exposure and use. At the beginning of each assessment, participants and their guardians are assented and consented, respectively, to the protocol elements for that year.

## Measures

### Outcomes: Neurocognitive Performance

Neurocognitive tasks are administered by research assistants (RAs) via iPads. NIH Toolbox includes multiple tasks to assess cognition, five of which were used as outcome variables because they were given at the same waves of data collection as SCS questions. Uncorrected standard scores are used for all measures to better assess within-person change. The following cognitive tasks are included in this analysis: ***Picture Sequence*** measures episodic memory. A series of images are shown to participants who are then asked to order the images in the order they viewed them. Participants complete two rounds of this task with the second task including additional images. ***Picture Vocabulary*** measures receptive vocabulary. Participants are instructed to select one of four presented images that best represents the provided word. ***Pattern Comparison Processing Speed*** measures processing. Participants were presented with two images per trial and prompted to respond with whether or not the images are identical. ***Oral Reading Recognition*** measures reading decoding skills and estimates crystalized abilities. In this task, participants are asked to pronounce letters and words that appear on the screen. ***Flanker*** measures inhibitory control and attention. During this task, participants are presented with a row of arrows and asked to focus on the middle arrow to select the direction it is pointing in, regardless of the direction the surrounding arrows are pointing.

### Predictors and Covariates

#### Self-reported Substance Use

Each year participants completed an RA-led substance use interview ^28^. Participants were first read a list of substances and asked if they used any in the past year, or since their last visit. RAs then completed a Timeline Follow-Back (TLFB) with daily substance use since their last visit for those who endorsed more than a sip/puff of any substance (alcohol, cannabis, etc.). Of particular interest here, binary (yes/no) past-year alcohol use (more than a sip), past-year nicotine use (more than a puff), and past-year other (non-cannabis) substance use are included as covariates in cognitive models. Lifetime report of personal cannabis use (either a puff/taste or more) were also reported for prevalence estimates (Aim 1) and exclusion in matching processes (Aim 2; see Statistical Analysis below).

#### Toxicological testing

Substance use was also assessed from biospecimen collection including hair, oral fluid, and urine ^29^. Hair collection was done for all participants who agreed at all timepoints. In a subset of participants with hair collected, toxicological testing identified metabolites for cannabis, nicotine, and alcohol (^30^). A full drug urinalysis was completed for all participants beginning at Year 4. Of interest here, any indicator of cannabis use across toxicological measures was used combined with self-report so that either was considered indicative of personal cannabis use.

#### Prenatal exposure

At Baseline, caregivers provided a retrospective self-report of prenatal substance use. Informants were asked about the mother’s use of cannabis and other substances before and after learning about the pregnancy. Binary prenatal exposure variables by substance were created, with separate reporting of alcohol, nicotine, marijuana, and any other illicit drug exposure. Single binary variables accounting for use of each drug class at any point during pregnancy were created for matching.

#### Parent History of AUD/SUD

Parent history of problems related to alcohol and/or substance use were queried at Baseline. Informants who had knowledge of the child’s biological relatives were asked if biological family members ever had issues due to substance use (e.g., alcohol/substance use–related separation/divorce, being laid off/fired related to alcohol/substance use problems, arrests/driving under the influence, being suspended or expelled from school 2 or more times, alcohol/substance harming health, being in an alcohol/substance treatment program, and causing arguments or being drunk/intoxicated a lot). They were then asked to indicate which family members experienced which issues. For this study, any report of biological father and biological mother problems related to AUD or, separately, SUD was coded for matching.

#### Sociodemographics

Guardians reported sociodemographic factors such as sex, household income, race/ethnicity, socioeconomic status, and age at Baseline ^31^. Sociodemographic factors were included in matching given prior research indicating these variables, including race/ethnicity, attenuate SCS-cognition associations ^22^.

#### SCS Exposure and Coding

Beginning at Year 2, caregivers reported whether their child had been exposed to SCS in the home or in a vehicle. They are asked similar questions about STS. Across waves, this was queried five times (Years 2-6). For creating time-varying predictor variables, this response was coded two ways: first, once reported, the first wave where SCS was reported, and every future visit was scored as 1 (regardless of whether exposure was reported again in follow-up visits). Secondarily, to assess repeat exposures over time, number of waves of reported exposure were summed across time (range=0-5, across all waves of data collection). Exposure to STS was also calculated using similar methods for prevalence estimates and to use in matching.

### Statistical Analyses

All participants with any data through the Year 6 Follow-Up were considered for inclusion. As SCS data was not collected at Baseline or Year 1, nor was broad-based neurocognitive testing conducted at Years 3 or 5, Aim 2 analyses were restricted to Years 2, 4, and 6, with SCS exposure at Year 3 and 5 collapsed into the following annual visit (Year 4 and 6, respectively). R version 4.5.0 ^32^ via RStudio ^33^ was employed for all statistical analyses.

#### Descriptives and Prevalence Estimates

Sociodemographics were examined via chi-square tests and are included in Table 1. Group differences by SCS exposure are described.

**Table 1.** Sociodemographic and Substance Exposure Characteristics of Matched Participants included in Cognitive Models.

| **Variable**<br>**Mean (SD)**<br>**or %** | **Controls**<br>**N=838** | **SCS**<br>**N=419** | **Sig. Dif.** |
| --- | --- | --- | --- |
| Age at Baseline | 9.88 (.62) | 9.88 (.61) | $p = .24$ |
| Female | 47% | 47% | $p = .99$ |
| Caregiver Education | | | $p = .99$ |
| <High School | 2% | 2% |  |
| High School/GED | 13% | 13% |  |
| Some College | 29% | 33% |  |
| Bachelor's | 29% | 26% |  |
| Post-Grad | 28% | 27% |  |
| Household Income | | | $p = .88$ |
| <25K | 15% | 18% |  |
| ≥25K & <50K | 17% | 17% |  |
| ≥50K & <75K | 18% | 17% |  |
| ≥75K & <100K | 11% | 12% |  |
| ≥100K & <200K | 27% | 25% |  |
| ≥200K | 5% | 5% |  |
| Race/Ethnicity | | | $p < .96$ |
| Asian | <1% | <1% |  |
| Black | 21% | 23% |  |
| Hispanic | 12% | 11% |  |
| White | 52% | 52% |  |
| Other | 15% | 14% |  |
| Secondhand Tobacco Exposure | 48.4% | 48.4% | $p = .99$ |
| Prenatal Alcohol Exposure | 33.2% | 33.9% | $p = .85$ |
| Prenatal Nicotine Exposure | 24.7 | 27.9% | $p = .24$ |
| Prenatal Cannabis Exposure | 15.5% | 21% | $p = .02$ |
| Prenatal Other Drug Exposure | 2% | <2% | $p = .83$ |
| Parental SUD |  |  |  |
| Mother | 4.2% | 5.5% | $p < .40$ |
| Father | 15% | 14.8% | $p = .99$ |
| Parental AUD |  |  |  |
| Mother | 7.5% | 9.1% | $p = .37$ |
| Father | 21.4% | 19.3% | $p = .45$ |
| Lifetime |  |  |  |
| Alcohol use | <2% | 0% | $p = .54$ |
| Nicotine Use | 6.7% | 7.4% | $p = .72$ |
| Other Drug Use | <2% | <4% | $p = .57$ |
| Number of Visits Reporting SCS |  |  |  |
| 1 Visits | -- | 58.5% |  |
| 2 Visits | -- | 21% |  |
| 3 Visits | -- | 10.3% |  |
| 4 Visits | -- | 6.2% |  |
| 5 Visits | -- | 4.1% |  |

Notes: Caregiver education refers to the highest level of education attained by primary caregivers; SUD=Substance Use Disorder; AUD=Alcohol Use Disorder; SCS=secondhand cannabis smoke

For Aim 1, in order to estimate national prevalence of SCS in U.S. adolescents, inverse propensity weights were applied to better reflect U.S. demography. The ABCD study provides weights based on Baseline sociodemographic and recruitment information to reflect national representation for 9-10 year-olds ^34^. We used the *survey* package in R ^35^ to apply these weights to the current dataset.

For prevalence of personal cannabis use, rates of use were compared descriptively between those with SCS and those without, with a simple logistic regression with no covariates assessing whether SCS exposure predicted lifetime cannabis use status by 17.

#### Data Cleaning Prior to Matching and Cognitive Models for Aim 2

Missingness. In order to match participants, complete cases on matching variables were required. Missingness was addressed utilizing multivariate imputation by chained equations in the MICE package ^36^ using 50 imputation iterations per variable. Variables with missingness imputed included parental education, family income, and race/ethnicity, each <1%.

#### Participant Selection and Matching

To solely examine associations between secondhand cannabis exposure and cognition, any indication of personal use was exclusionary from matching. Therefore, participants who self-reported cannabis use at any point throughout the study and/or had toxicological hair or urine data indicative of presence of metabolized cannabinoids (i.e., THCCOOH, which shows they have personally ingested cannabis^37^) were excluded. Of the remaining participants (n=419 with SCS), MatchIt was used to match the exposed group to similar participants who had not been exposed to secondhand cannabis smoke nor used cannabis personally (by self-report or toxicological testing). Specifically, participants were matched on sociodemographic characteristics (parental education, household income, self-identified race and ethnicity, age at Baseline, sex), ever reported exposure to secondhand nicotine (cigarettes, vaping, or other), prenatal substance use exposure, and parental history of AUD and SUD. Given the large sample size of the full cohort, a 2:1 match ratio was selected (2 Controls:1 Treated). Match diagnostics demonstrated good balance with standard mean difference (SMD) below 0.1 for all variables, with the exception of prenatal cannabis exposure (SMD=.135). While an SMD < .25 is generally viewed as acceptable ^38^, in order to ensure doubly robust methods, sensitivity analyses were conducted including variables with SMD > .1 (i.e., prenatal cannabis exposure) in primary cognitive outcome model; as results were consistent across models, sensitivity analyses are included within the Supplement only for the significant models.

#### Linear mixed effects (LME)

Models were run using *lme4* ^39^ to examine the influence of SCS exposure on neurocognitive performance over time (Year 2, 4, 6 Follow-Up). Separate LMEs were modeled for each of the five neurocognitive task, with time-varying SCS status over the 5 year testing period*age as the primary predictor of interest. Given robust controls through matching (sociodemographic, prenatal exposure; secondhand tobacco exposure) and exclusion criteria (no personal cannabis use), covariates were limited to time-varying (self-reported past-year binary alcohol use, nicotine use, other drug use) and other relevant factors (i.e., sex), while nesting within subject and family. Inclusion of random slopes of participant ID did not significantly improve fit (for the primary Flanker model, χ²(2) = 1.60, p = 0.45), and therefore models with random intercepts only were retained. In secondary models, for neurocognitive tasks significant in primary models, time-varying cumulative number of years of SCS exposure (0- 5)*age was the primary predictor of interest with all other variables the same. Given the pattern of results, quadratic modeling was examined; as fit was not improved, the linear model was retained and is reported below.

Results were considered significant if *p*<.05. Standardized β were calculated using the *MuMin* package ^40^, acknowledging that ABCD is powered to detect very small effect sizes ^41^. Estimated marginal means to test group differences at each age integer were extracted via the *emmeans* package ^42^ and pairwise tested. Results include 95% Wald confidence intervals.

## Results

### Aim 1. Secondhand Cannabis Smoke Exposure Prevalence and Personal Cannabis Use

#### Participant Characteristics

Across the full ABCD cohort through Year 6 Follow-Up, 776 (6.9%) participants ever reported secondhand cannabis exposure; in contrast, 1,637 (14.5%) ever reported secondhand nicotine exposure. A small portion of the full cohort (n=434, 3.8%) reported exposure to both secondhand cannabis and secondhand nicotine. Propensity weighted estimates indicate general U.S. prevalence of in home SCS exposure at ages 11-17 is 7.3% (95%CI:6.7-7.8%). Of those with SCS (n=776), 357 were identified as having lifetime personal cannabis use by Year 6 (and thus were excluded from further analyses on cognition). Youth with SCS exposure were significantly more likely to report lifetime cannabis use by Year 6 (OR=3.83[95%CI:3,29,4.44], b=1.34, p<.0001); 46% reported lifetime cannabis use by Year 6, relative to only 20% of lifetime cannabis use among non-SCS exposed youth in ABCD.

### Aim 2. Secondhand Cannabis Smoke Exposure without Personal Cannabis Use

#### Sociodemographics of Matched Participants

Table 1 displays sociodemographic and other participant characteristics for those used in longitudinal cognitive modeling. Included participants were, on average, 9.9 years-old at ABCD enrollment, and between ages 10.6-17.8 within the present data analyses. As expected with matching, groups did not differ significantly on sociodemographic factors or matching criteria. Youth in the SCS group did not differ on self-reported lifetime substance use (*p*’s > .5). However, youth in the SCS exposure group did have higher rates of prenatal cannabis exposure (p=.02).

### Neurocognition Related to SCS Exposure without Personal Cannabis Use

#### Primary Neurocognitive Models

Full models are presented in the **Table 2** and **Tables S1-S4** for non-significant models, with a primary interaction of SCS and age reported here. For attention and inhibitory control (Flanker Task; n=1,165; n_obs_=2,656), SCS exhibited a main effect (β=0.29, b=5.91, 95%CI:0.22,11.61, *p*=0.042) as did age (β=-0.36, b=-0.42, 95%CI:1.47-1.77, *p*<.001). Further, SCS*age demonstrated a significant interaction (β=-0.32, b=-0.42, 95%CI:- 0.80,-0.05, *p*=.028; see **Figure 1**). Review of estimated marginal means indicate no group difference at ages 10-15, a marginal difference at age 16 (p=.057), and a significant difference at age 17 where the unexposed group demonstrated better performance than the SCS group at age 17 (*p*=.02; see **Table S5**).

**Figure 1.**
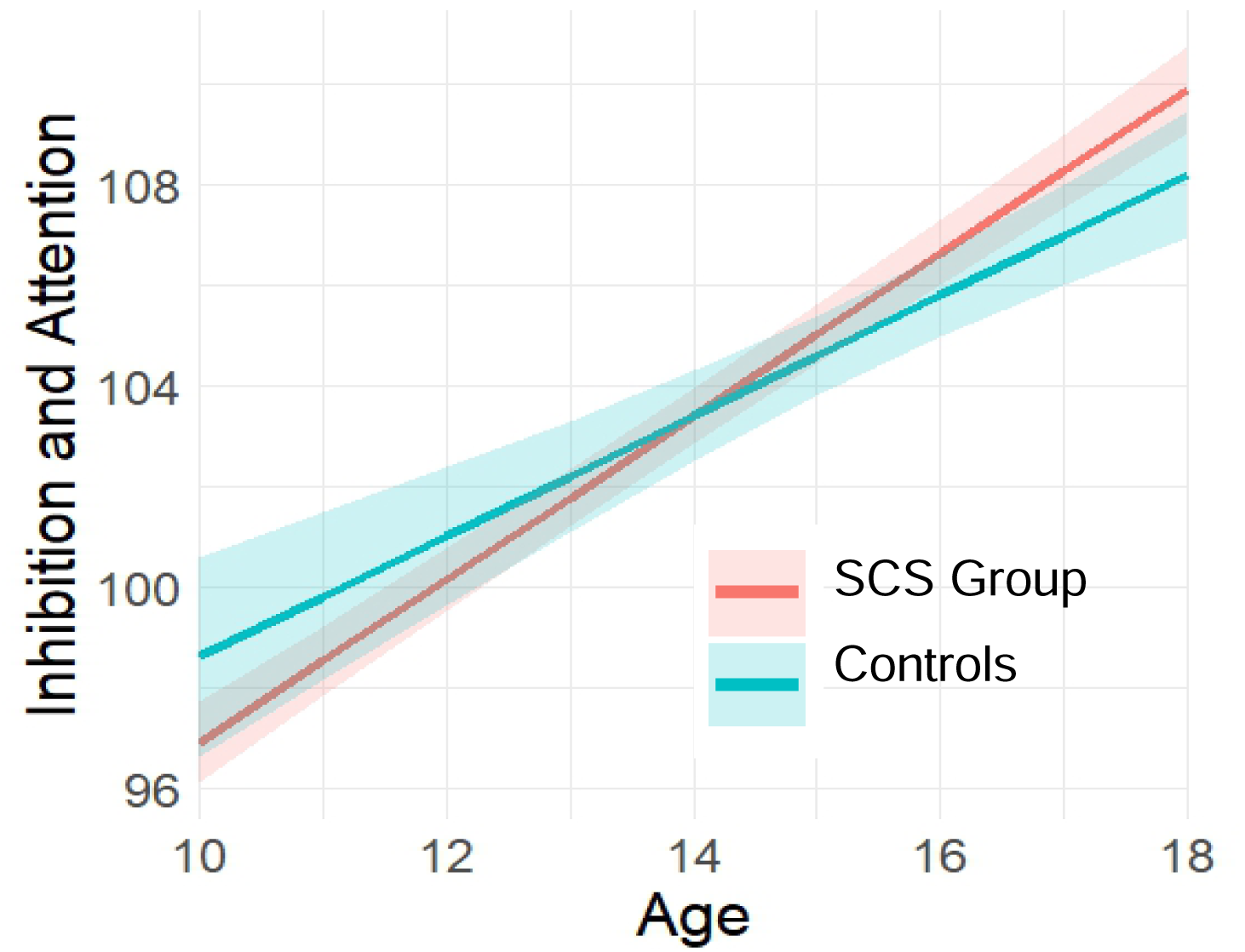
Interaction between SCS group status and age on Flanker Inhibitory Control and Attention task performance Notes: SCS Group = time-varying secondhand cannabis exposure, such that once a participant is in the SCS group, they continue in that group

**Table 2.**
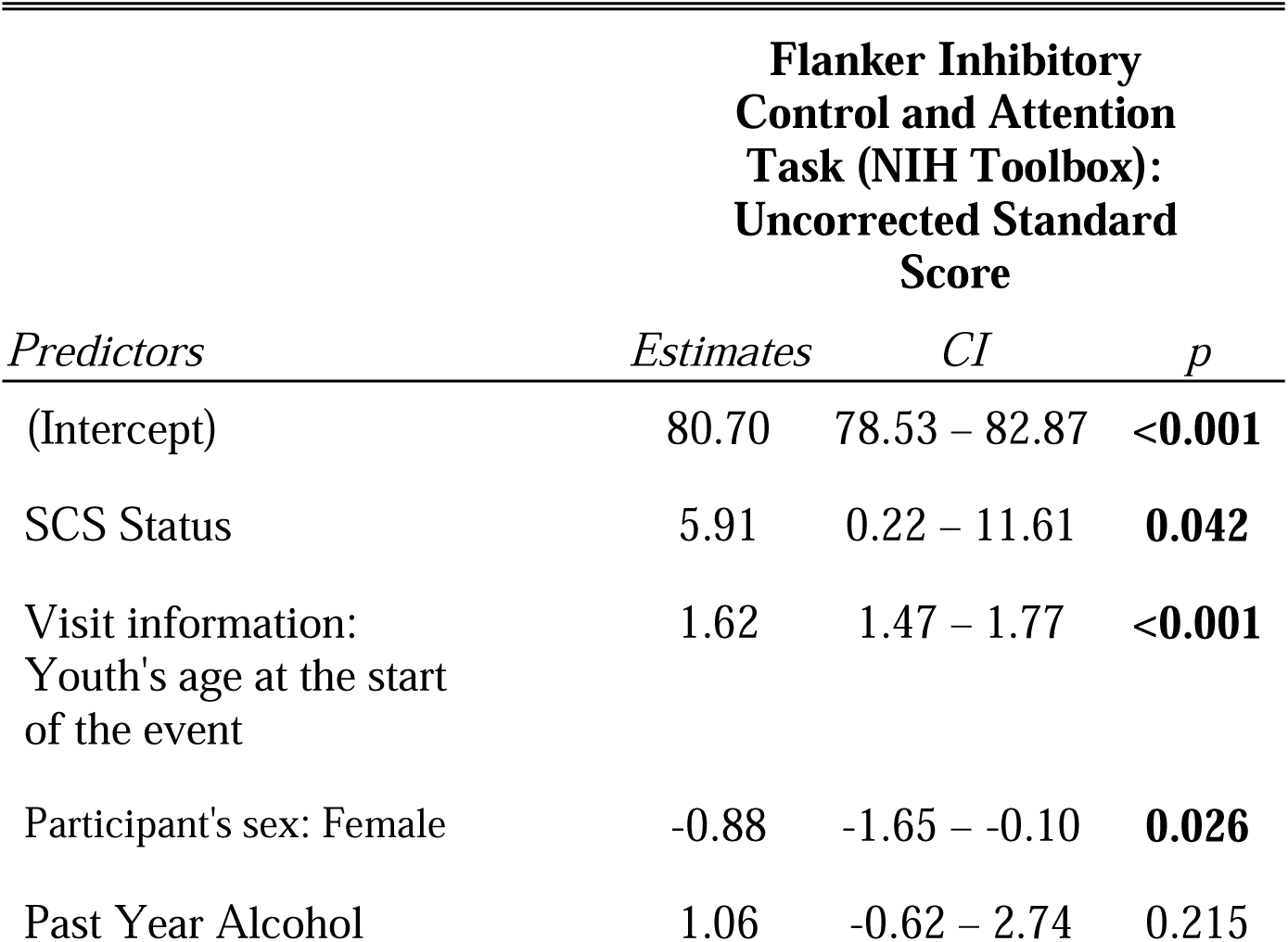
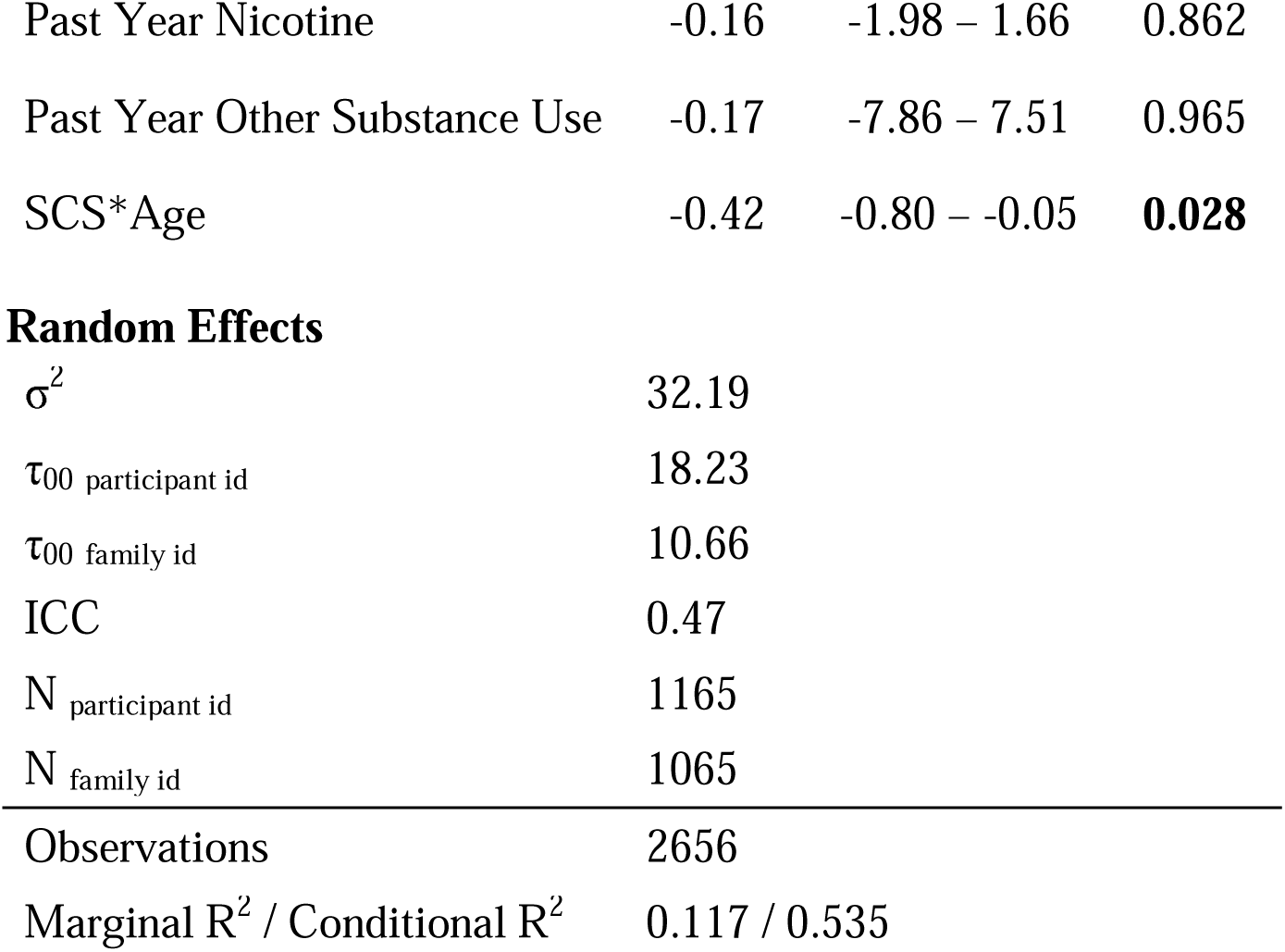
.Primary Neurocognitive Models Full Output—Flanker Inhibitory Control and Attention Task.

| *Predictors* | **Flanker Inhibitory**<br>**Control and Attention**<br>**Task (NIH Toolbox):**<br>**Uncorrected Standard**<br>**Score** |  |  |
| --- | --- | --- | --- |
|  | *Estimates* | *CI* | *p* |
| (Intercept) | 80.70 | 78.53 – 82.87 | **<0.001** |
| SCS Status | 5.91 | 0.22 – 11.61 | **0.042** |
| Visit information:<br>Youth's age at the start<br>of the event | 1.62 | 1.47 – 1.77 | **<0.001** |
| Participant's sex: Female | -0.88 | -1.65 – -0.10 | **0.026** |
| Past Year Alcohol | 1.06 | -0.62 – 2.74 | 0.215 |
| Past Year Nicotine | -0.16 | -1.98 – 1.66 | 0.862 |
| Past Year Other Substance Use | -0.17 | -7.86 – 7.51 | 0.965 |
| SCS*Age | -0.42 | -0.80 – -0.05 | **0.028** |

Random Effects
|  |  |
| --- | --- |
| $\sigma^2$ | 32.19 |
| $\tau_{00}$ participant id | 18.23 |
| $\tau_{00}$ family id | 10.66 |
| ICC | 0.47 |
| $N_{\text{participant id}}$ | 1165 |
| $N_{\text{family id}}$ | 1065 |
| Observations | 2656 |
| Marginal $R^2$ / Conditional $R^2$ | 0.117 / 0.535 |

In a sensitivity analysis, SCS*age results remained significant (β=-0.34, b=-0.45, 95%CI:-0.83,-0.07, *p*=019) when adding prenatal cannabis exposure as a covariate to confirm results despite imperfect match (see **Table S6**).

No other models demonstrated significant associations between SCS group status moderated by age on neurocognitive performance.

#### Secondary Neurocognitive Models

Number of years of SCS exposure marginally interacted with age in predicting Flanker performance, with fewer exposures being associated with better performance over time (β=-0.39, b=-0.25, 95%CI:-0.52,-0.001, *p*=0.057; see **Figure 2** and **Table S7**).

**Figure 2.**
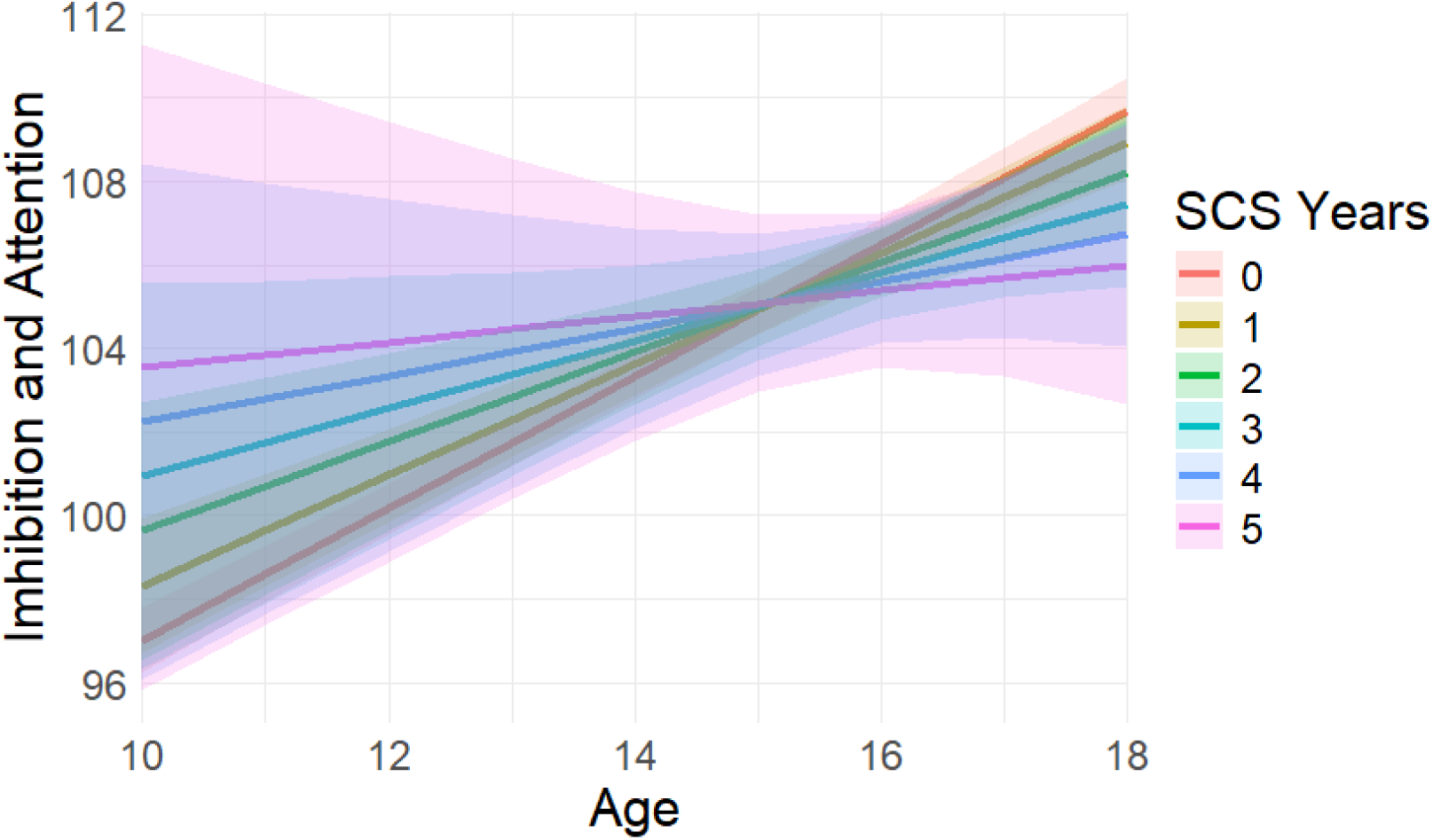
Interaction between number of waves of SCS exposure and age on Flanker Inhibitory Control and Attention task performance Notes: SCS Years = number of cumulative waves (follow-up years) of secondhand cannabis (SCS) exposure

## Discussion

This study aimed to (1) explore the prevalence of adolescents with SCS and those who also used cannabis, and (2) examine cognitive trajectories of those exposed to SCS, but who did not use cannabis themselves. Results indicate that 46% of those who reported exposure to SCS also used cannabis themselves by age 17, nearly twice the rate of peers without SCS exposure (20%). Furthermore, adolescents with SCS exposure demonstrated reduced improvement over time in attention and inhibition relative to their non-exposed peers, with marginal interaction between number of visits with reported exposure and performance such that fewer reported exposures were related to better performance on an attention/inhibition task over time. It is notable that these results were of medium effect size and robust to significant controls, including a well-matched non-exposed group, excluding for personal cannabis use, and covarying for other substance use. On balance, no other cognitive domains demonstrated differences by exposure status, suggesting SCS may have domain-specific relationships with attention and inhibitory control.

As prevalence of cannabis use is generally increasing among U.S. adults ^4–6^, corresponding increases in exposure to SCS for youth may be expected. Here, estimated prevalence of SCS exposure in adolescents is around 7%. One factor contributing to the increase of in-home SCS by parents may be the growing perception that SCS exposure is less harmful than STS exposure. A nationwide survey conducted at two timepoints indicated that 35% of respondents agreed that SCS exposure is less harmful than STS exposure in 2017, and 40% agreed in 2021 ^43^. Additionally, in-home smoke is less likely among those who perceive SCS as more harmful ^44^, suggesting a potential association between perceived harm and likelihood of smoking cannabis in-home. Future research is needed to clarify harms associated with SCS, with subsequent public health efforts to inform families.

Data here reveals that nearly half of the adolescents who were exposed to SCS also used cannabis themselves, with 3.8 times increase in odds of personal use relative to youth without SCS. This is consistent with Social Learning Theory ^45^, in which social context (e.g. family, friends, environment) influences cognition and behavior. Familial cannabis use, while youth are present, may influence adolescents to use themselves. Furthermore, there is a growing body of research suggesting familial cannabis use is associated with earlier age of cannabis use onset and more frequent use ^9–11^. Given increases of in-home SCS with minors present, it will be important to further explore the interaction between social learning and substance use behavior in adolescents and young adults.

Primary cognitive trajectory analyses here demonstrate a relationship between SCS exposure and attention/inhibitory control over time, as those with SCS exposure showed reduced growth in performance from ages 10-17. Further, secondary analyses indicated a marginal effect of number of waves of reported exposure correlating with further plateaued trajectories, though the exact amount of exposure is uncertain. While the cause of this relationship remains unknown, it is plausible that the chemical makeup of the cannabis smoke is a driving factor. Trajectory profiles demonstrated in SCS exposed youth match results from adolescents with personal use (i.e., reduced improvement over time with better performance in the unexposed group by age 17) ^46^; however, personal use followed this pattern across a broad swath of cognitive domains while SCS-exposure was linked only to attention and inhibitory control. Importantly, participants in the present study with evidence of personal cannabis use were excluded from assessment of cognition associations. Considering evidence of carcinogens, toxins, and particulate matter in SCS ^17,18^, it may be more likely that cannabis smoke contaminants are contributing to downstream behavioral outcomes such as reduced improvement in attention over time rather than being due to THC or other cannabinoid constituents.

It is worth considering why SCS may relate specifically to attention and inhibitory control. Given SCS may contain greater particulate matter and contaminants than STS ^18–20^, it is possible that a similar mechanism is at work with parallels in the relationship to cognitive functioning. Evidence suggests particulate matter and contaminants from tobacco smoke can increase neuroinflammation, disrupt neural connectivity, and contribute to aberrant neurodevelopment ^47,48^. STS also increases carbon monoxide exposure and reduces blood oxygenation, which may have downstream behavior effects ^49^. Further, recent research indicates that early to mid-childhood youth exposed to STS similarly had decreased performance on the same attention and inhibitory control task as well as other executive function tasks, even after controlling for prenatal exposure ^50^. Yet SCS has not yet been investigated in a similar fashion. Thus, more mechanistic studies of SCS are needed. Research measuring levels and frequency of in-home SCS exposure and cognitive relationships would be beneficial in determining public health risks.

### Strengths, Limitations, and Future Directions

The present study is subject to both strengths and limitations. The ABCD Study obtains a large, diverse sample with access to both toxicology reports and self-report substance use measures. Additionally, a robust cognitive battery and clinical assessment are administered every other year, allowing for a wide range of cognitive domains to be analyzed. Statistical controls here were robust, with thorough matching (including on prenatal exposure, STS, and sociodemographics), excluding personal use in cognition analyses, and covarying for other substance use. Although within-subject trajectories were measured, causation cannot be inferred due to the observational nature of this study. This study is one of the first known studies to analyze the interaction between age and cognitive trajectories within adolescents, but the age of first SCS exposure is not queried. Future study designs could consider cognitive testing prior to SCS exposure to test causal influences of exposure and impacts at specific ages. Personal cannabis use was exclusionary for modeling, with use identified both through toxicological testing and self-report; in extreme circumstances, urine can be positive from SCS alone ^51^. SCS was only assessed within the home and family vehicles, but it is possible that adolescents are also being exposed to cannabis outside of the home (e.g. with friends, with non-residing relatives, at school, at work), which should continue to be explored in future work. Additionally, though toxicology reports provide great insight to cannabis exposure and use patterns, these measures only capture a few days up to a 3-month window of detection which does not reflect the full period between assessments. While sociodemographic factors were accounted for within this study, it should be noted that marginalized populations may be particularly susceptible to SCS exposure: in-home familial cannabis smoke with minors present is more common among single parent households, and among multi-unit housing, both of which are indicators of socioeconomic hardships ^5,52,53^. Public health initiatives should continue to address systemic issues that can cause health disparities due to increased exposure to SCS among marginalized populations.

### Conclusions

Research into SCS exposure is in its nascent stages, despite cannabis policy changes and increases in personal use and SCS prevalence in youth. In the first known longitudinal analyses of SCS-exposed adolescents, data here indicates adolescents with SCS exposure have over twice the rate of personal cannabis use and, in those who do not use cannabis themselves, demonstrate reduced improvement in attention and inhibitory control over time. This is even after robust sociodemographic controls through matching. Cognitive findings may reflect a similar mechanism as secondhand tobacco exposure, as STS exposed youth similarly exhibit impaired attention. More research is necessary regarding the basic science of contaminants in SCS and clinical studies of youth with exposure, including with objective measures of exposure. Parallels between tobacco and cannabis suggest that similar public health frameworks may be necessary ^54^, and encourage expediting translation of scientific findings into actionable public health initiatives ^55^. Public health and policymakers should make efforts to curb youth SCS exposure, given the potential for risk which has not been fully explored to date.

## Data Availability

All data is available to approved researchers via https://www.nbdc-datahub.org/

## Acknowledgements

Authors would like to thank the families who participate in the ABCD Study and the staff who collect the data. Data used in the preparation of this article were obtained from the <u>Adolescent Brain Cognitive Development™ (ABCD) Study</u>, held in the <u>NIH Brain Development Cohorts Data Sharing Platform</u>. This is a multisite, longitudinal study designed to recruit more than 10,000 children aged 9–10 and follow them over 10 years into early adulthood. ABCD Consortium investigators designed and implemented the study and/or provided data but did not necessarily participate in the analysis or writing of this report. This manuscript reflects the views of the authors and may not reflect the opinions or views of the NIH or ABCD Consortium investigators. The ABCD data repository grows and changes over time. The ABCD data used in this report came from DOI 10.1.66.93/8f3w-5260. Authors can be contacted for analytical code.

## References

1. Zerleen S. Quader, P.; Douglas R. Roehler, P.; Alana M. Vivolo-Kantor, P.; Jean Y. Ko, P. Routes of Marijuana Use — Behavioral Risk Factor Surveillance System, 22 U.S. States and Two Territories, 2022. MMWR Morb. Mortal. Wkly. Rep. 2025, 74. 10.15585/mmwr.mm7412a1.

2. Steigerwald, S.; Wong, P. O.; Cohen, B. E.; Ishida, J. H.; Vali, M.; Madden, E.; Keyhani, S. Smoking, Vaping, and Use of Edibles and Other Forms of Marijuana Among U.S. Adults. Ann. Intern. Med. 2018, 169 (12), 890–892. 10.7326/M18-1681.

3. Tripathi, O.; Posis, A. I. B.; Thompson, C. A.; Ferris, J.; Anuskiewicz, B.; Nguyen, B.; Liles, S.; Berardi, V.; Zhu, S.-H.; Winstock, A.; Bellettiere, J. In-Home Cannabis Smoking Among a Cannabis-Using Convenience Sample from the Global Drug Survey: With Weighted Estimates for U.S. Respondents. Cannabis Cannabinoid Res. 2024, 9 (1), 353–362. 10.1089/can.2022.0139.

4. Bellettiere, J. In-Home Cannabis Smoking More Prevalent than in-Home Tobacco Smoking among 2019 Global Drug Survey Respondents. 2022. 10.1016/j.addbeh.2021.107130.

5. Goodwin, R. D.; Cheslack-Postava, K.; Santoscoy, S.; Bakoyiannis, N.; Hasin, D. S.; Collins, B. N.; Lepore, S. J.; Wall, M. M. Trends in Cannabis and Cigarette Use Among Parents With Children at Home: 2002 to 2015. Pediatrics 2018, 141 (6), e20173506. 10.1542/peds.2017-3506.

6. Goodwin, R. D.; Kim, J. H.; Cheslack-Postava, K.; Weinberger, A. H.; Wu, M.; Wyka, K.; Kattan, M. Trends in Cannabis Use among Adults with Children in the Home in the United States, 2004-2017: Impact of State-Level Legalization for Recreational and Medical Use. Addiction 2021, 116 (10), 2770–2778. 10.1111/add.15472.

7. Tripathi, O.; Bellettiere, J.; Liles, S.; Shi, Y. Location and Home Rules of Cannabis Use – Findings from Marijuana Use and Environmental Survey 2020, a Nationally Representative Survey in the United States. Prev. Med. Rep. 2023, 35, 102289. 10.1016/j.pmedr.2023.102289.

8. Shorey, R. C.; Fite, P. J.; Elkins, S. R.; Frissell, K. C.; Tortolero, S. R.; Stuart, G. L.; Temple, J. R. The Association Between Problematic Parental Substance Use and Adolescent Substance Use in an Ethnically Diverse Sample of 9th and 10th Graders. J. Prim. Prev. 2013, 34 (6), 381–393. 10.1007/s10935-013-0326-z.

9. Kerr, D. C. R.; Tiberio, S. S.; Capaldi, D. M. Contextual Risks Linking Parents’ Adolescent Marijuana Use to Offspring Onset. Drug Alcohol Depend. 2015, 154, 222–228. 10.1016/j.drugalcdep.2015.06.041.

10. Cui, Y.; Wang, Y.; LoParco, C. R.; Romm, K. F.; Cavazos-Rehg, P. A.; Chakraborty, R.; McCready, D. M.; Yang, Y. T.; Berg, C. J. Indicators of Intergenerational Transmission of Cannabis Use Among US Young Adults. Subst. Use Addict. J. 2025, 46 (4), 960–971. 10.1177/29767342251337212.

11. O’Loughlin, J. L.; Dugas, E. N.; O’Loughlin, E. K.; Winickoff, J. P.; Montreuil, A.; Wellman, R. J.; Sylvestre, M.-P.; Hanusaik, N. Parental Cannabis Use Is Associated with Cannabis Initiation and Use in Offspring. J. Pediatr. 2019, 206, 142–147.e1. 10.1016/j.jpeds.2018.10.057.

12. Bailey, J. A.; Epstein, M.; Steeger, C. M.; Hill, K. G. Concurrent and Prospective Associations Between Substance-Specific Parenting Practices and Child Cigarette, Alcohol, and Marijuana Use. J. Adolesc. Health 2018, 62 (6), 681–687. 10.1016/j.jadohealth.2017.11.290.

13. Gray, K. M.; Squeglia, L. M. Research Review: What Have We Learned About Adolescent Substance Use? J. Child Psychol. Psychiatry 2018, 59 (6), 618–627. 10.1111/jcpp.12783.

14. Muheriwa-Matemba, S. R.; Baral, A.; Abdshah, A.; Diggs, B.-N. A.; Gerber Collazos, K. S.; Morris, K. B.; Messiah, S. E.; Vidot, D. C. Cardiovascular and Respiratory Effects of Cannabis Use by Route of Administration: A Systematic Review. Subst. Use Misuse 2024, 59 (9), 1331–1351. 10.1080/10826084.2024.2341317.

15. Gorelick, D. A. Cannabis-Related Disorders and Toxic Effects. N. Engl. J. Med. 2023, 389 (24), 2267–2275. 10.1056/NEJMra2212152.

16. Chandy, M.; Nishiga, M.; Wei, T.; Nadeau, K.; Wu, J. C. Adverse Impact of Marijuana on Human Health. Annu. Rev. Med. 2024, 75, 353–367. 10.1146/annurev-med-052422-020627.

17. Moir, D.; Rickert, W. S.; Levasseur, G.; Larose, Y.; Maertens, R.; White, P.; Desjardins, S. A Comparison of Mainstream and Sidestream Marijuana and Tobacco Cigarette Smoke Produced under Two Machine Smoking Conditions. Chem. Res. Toxicol. 2008, 21 (2), 494–502. 10.1021/tx700275p.

18. Janssen, F. Comparison Between Smoked Tobacco and Medical Cannabis Cigarettes Concerning Particulate Matter. 2024, 9 (6), 1492–1499. 10.1089/can.2023.0201.

19. Ott, W. R.; Wallace, L. A.; Cheng, K.-C.; Hildemann, L. M. Measuring PM2.5 Concentrations from Secondhand Tobacco vs. Marijuana Smoke in 9 Rooms of a Detached 2-Story House. Sci. Total Environ. 2022, 852, 158244. 10.1016/j.scitotenv.2022.158244.

20. Nguyen, P. K.; Hammond, S. K. Fine Particulate Matter Exposure From Secondhand Cannabis Bong Smoking. *JAMA Netw*. Open 2022, 5 (3), e224744. 10.1001/jamanetworkopen.2022.4744.

21. Zhou, S.; Rosenthal, D. G.; Sherman, S.; Zelikoff, J.; Gordon, T.; Weitzman, M. Physical, Behavioral, and Cognitive Effects of Prenatal Tobacco and Postnatal Secondhand Smoke Exposure. Curr. Probl. Pediatr. Adolesc. Health Care 2014, 44 (8), 219–241. 10.1016/j.cppeds.2014.03.007.

22. Wade, N. E. Clouding up Cognition? Secondhand Cannabis and Tobacco Exposure Related to Cognitive Functioning in Youth. 2022. 10.1016/j.bpsgos.2022.01.010.

23. Moore, B. Associations between Prenatal and Postnatal Exposure to Cannabis with Cognition and Behavior at Age 5 Years: The Healthy Start Study. 2023. 10.3390/ijerph20064880.

24. Satybaldiyeva, N. Secondhand Smoke Exposure and Asthma Status among Adolescents: Findings from the 2019-2020 California Student Tobacco Survey. 2024. 10.1016/j.pmedr.2024.102842.

25. Cohn, A. Secondhand Cannabis Smoke Exposure and Respiratory Symptoms among Adults Living in a State with Legalized Medical Cannabis with Limited Smoke-Free Protections. 2024. 10.1016/j.pmedr.2024.102835.

26. Jernigan, T. L.; Brown, S. A. Introduction. Dev. Cogn. Neurosci. 2018, 32, 1–3. 10.1016/j.dcn.2018.02.002.

27. Casey, B. J.; Cannonier, T.; Conley, M. I.; Cohen, A. O.; Barch, D. M.; Heitzeg, M. M.; Soules, M. E.; Teslovich, T.; Dellarco, D. V.; Garavan, H.; Orr, C. A.; Wager, T. D.; Banich, M. T.; Speer, N. K.; Sutherland, M. T.; Riedel, M. C.; Dick, A. S.; Bjork, J. M.; Thomas, K. M.; Chaarani, B.; Mejia, M. H.; Hagler, D. J.; Daniela Cornejo, M.; Sicat, C. S.; Harms, M. P.; Dosenbach, N. U. F.; Rosenberg, M.; Earl, E.; Bartsch, H.; Watts, R.; Polimeni, J. R.; Kuperman, J. M.; Fair, D. A.; Dale, A. M. The Adolescent Brain Cognitive Development (ABCD) Study: Imaging Acquisition across 21 Sites. Dev. Cogn. Neurosci. 2018, 32, 43–54. 10.1016/j.dcn.2018.03.001.

28. Lisdahl, K. M.; Sher, K. J.; Conway, K. P.; Gonzalez, R.; Feldstein Ewing, S. W.; Nixon, S. J.; Tapert, S.; Bartsch, H.; Goldstein, R. Z.; Heitzeg, M. Adolescent Brain Cognitive Development (ABCD) Study: Overview of Substance Use Assessment Methods. Dev. Cogn. Neurosci. 2018, 32, 80–96. 10.1016/j.dcn.2018.02.007.

29. Uban, K. A.; Horton, M. K.; Jacobus, J.; Heyser, C.; Thompson, W. K.; Tapert, S. F.; Madden, P. A. F.; Sowell, E. R. Biospecimens and the ABCD Study: Rationale, Methods of Collection, Measurement and Early Data. Dev. Cogn. Neurosci. 2018, 32, 97–106. 10.1016/j.dcn.2018.03.005.

30. Wade, N. E.; Si, Y.; Tapert, S. F.; Linkersdörfer, J.; Lisdahl, K. M.; Moore, H. R.; Tally, L.; Das, B.; Huestis, M. A.; Wallace, A. L.; Sullivan, R. M.; Szpak, V.; Zhang, L.; Ziemer, L.; Thompson, W. K. Weighted Prevalence of Biochemically Verified Substance Use in Healthy Adolescents across the USA. Addiction 2026. 10.1111/add.70548.

31. Barch, D. M.; Albaugh, M. D.; Avenevoli, S.; Chang, L.; Clark, D. B.; Glantz, M. D.; Hudziak, J. J.; Jernigan, T. L.; Tapert, S. F.; Yurgelun-Todd, D.; Alia-Klein, N.; Potter, A. S.; Paulus, M. P.; Prouty, D.; Zucker, R. A.; Sher, K. J. Demographic, Physical and Mental Health Assessments in the Adolescent Brain and Cognitive Development Study: Rationale and Description. Dev. Cogn. Neurosci. 2018, 32, 55–66. 10.1016/j.dcn.2017.10.010.

32. R Core Team. R: A Language and Environment for Statistical Computing. 2026.

33. Posit team. RStudio: Integrated Development Environment for R. 2025.

34. Heeringa, S. G.; Berglund, P. A. A Guide for Population-Based Analysis of the Adolescent Brain Cognitive Development (ABCD) Study Baseline Data. bioRxiv February 10, 2020, p 2020.02.10.942011. 10.1101/2020.02.10.942011.

35. Lumley, T. survey: analysis of complex survey samples. R-universe. 2026. 10.32614/CRAN.package.survey.

36. Buuren, S. van; Groothuis-Oudshoorn, K. Mice: Multivariate Imputation by Chained Equations in R. J. Stat. Softw. 2011, 45, 1–67. 10.18637/jss.v045.i03.

37. Ramirez Fernandez, M. D. M.; Cirimele, V.; Ruyssinckx, E.; Di Fazio, V.; Jacobs, W.; Schmit, G.; Wille, S. M. R. Challenges in the Interpretation of Isolated THC-COOH Detection in Hair: Forensic and Clinical Implications. Drug Test. Anal. 2026, 18 (6), 785–793. 10.1002/dta.70072.

38. Stuart, E. A.; Lee, B. K.; Leacy, F. P. Prognostic Score–Based Balance Measures Can Be a Useful Diagnostic for Propensity Score Methods in Comparative Effectiveness Research. J. Clin. Epidemiol. 2013, 66 (8, Supplement), S84–S90.e1. 10.1016/j.jclinepi.2013.01.013.

39. Bates, D.; Mächler, M.; Bolker, B.; Walker, S. Fitting Linear Mixed-Effects Models Using Lme4. J. Stat. Softw. 2015, 67, 1–48. 10.18637/jss.v067.i01.

40. Bartoń, K. MuMIn: Multi-Model Inference, 2026. 10.32614/CRAN.package.MuMIn.

41. Owens, M. M.; Potter, A.; Hyatt, C. S.; Albaugh, M.; Thompson, W. K.; Jernigan, T.; Yuan, D.; Hahn, S.; Allgaier, N.; Garavan, H. Recalibrating Expectations about Effect Size: A Multi-Method Survey of Effect Sizes in the ABCD Study. PloS One 2021, 16 (9), e0257535. 10.1371/journal.pone.0257535.

42. Lenth, R. V.; Piaskowski, J.; Banfai, B.; Bolker, B.; Buerkner, P.; Giné-Vázquez, I.; Hervé, M.; Jung, M.; Love, J.; Miguez, F.; Riebl, H.; Singmann, H.; Derumigny, A.; Schmidt, P. Emmeans: Estimated Marginal Means, Aka Least-Squares Means, 2026. 10.32614/CRAN.package.emmeans.

43. Chambers, J. Perceptions of Safety of Daily Cannabis vs Tobacco Smoking and Secondhand Smoke Exposure, 2017-2021. 2023. 10.1001/jamanetworkopen.2023.28691.

44. Tripathi, O. Clearing the Air: Heightened Perception of Harm from Secondhand Cannabis Smoke Exposure Is Associated with No in-Home Cannabis Smoking in a 21- Country Convenience Sample. 2024. 10.1016/j.ypmed.2024.108178.

45. Bandura, A. Social Learning Theory; Prentice-Hall: New Jersey, 1977.

46. Wade, N. E.; Sullivan, R. M.; Wallace, A. L.; Visontay, R.; Szpak, V.; Lisdahl, K. M.; Huestis, M. A.; Gonçalves, P. D.; Byrne, H.; Mewton, L.; Jacobus, J.; Tapert, S. F. Longitudinal Neurocognitive Trajectories in a Large Cohort of Youth Who Use Cannabis: Combining Self-Report and Toxicology. Neuropsychopharmacology 2026, 1–10. 10.1038/s41386-026-02395-1.

47. Rechtman, E.; Rebello, V.; Invernizzi, A.; Sather, A.; Oluyemi, K.; Rodriguez, M. A.; Horton, M. Environmental Exposures and Neuroimaging in Children with Neurodevelopmental Disorders: A Scoping Review. Curr. Environ. Health Rep. 2026, 13 (1), 20. 10.1007/s40572-026-00538-6.

48. Hajdusianek, W.; Żórawik, A.; Waliszewska-Prosół, M.; Poręba, R.; Gać, P. Tobacco and Nervous System Development and Function—New Findings 2015–2020. Brain Sci. 2021, 11 (6), 797. 10.3390/brainsci11060797.

49. Chen, R.; Clifford, A.; Lang, L.; Anstey, K. J. Is Exposure to Secondhand Smoke Associated with Cognitive Parameters of Children and Adolescents?-A Systematic Literature Review. Ann. Epidemiol. 2013, 23 (10), 652–661. 10.1016/j.annepidem.2013.07.001.

50. Fuemmeler, B. F.; Glasgow, T. E.; Schechter, J. C.; Maguire, R.; Sheng, Y.; Bidopia, T.; Barsell, D. J.; Ksinan, A.; Zhang, J.; Lin, Y.; Hoyo, C.; Murphy, S.; Qin, J.; Wang, X.; Kollins, S. Prenatal and Childhood Smoke Exposure Associations with Cognition, Language, and Attention-Deficit/Hyperactivity Disorder. J. Pediatr. 2023, 256, 77–84.e1. 10.1016/j.jpeds.2022.11.041.

51. Cone, E. J.; Bigelow, G. E.; Herrmann, E. S.; Mitchell, J. M.; LoDico, C.; Flegel, R.; Vandrey, R. Non-Smoker Exposure to Secondhand Cannabis Smoke. I. Urine Screening and Confirmation Results. J. Anal. Toxicol. 2015, 39 (1), 1–12. 10.1093/jat/bku116.

52. Chu, A. K.; Kaufman, P.; Chaiton, M. Prevalence of Involuntary Environmental Cannabis and Tobacco Smoke Exposure in Multi-Unit Housing. Int. J. Environ. Res. Public. Health 2019, 16 (18), 3332. 10.3390/ijerph16183332.

53. Repace, J. L. Secondhand Smoke Infiltration in Multiunit Housing: Health Effects and Nicotine Levels. Indoor Environ. 2024, 1 (2), 100013. 10.1016/j.indenv.2024.100013.

54. Barry, R. A.; Glantz, S. A Public Health Framework for Legalized Retail Marijuana Based on the US Experience: Avoiding a New Tobacco Industry. PLoS Med. 2016, 13 (9), e1002131. 10.1371/journal.pmed.1002131.

55. Harris, J. K.; Luke, D. A.; Zuckerman, R. B.; Shelton, S. C. Forty Years of Secondhand Smoke Research: The Gap Between Discovery and Delivery. Am. J. Prev. Med. 2009, 36 (6), 538–548. 10.1016/j.amepre.2009.01.039.

