## Supplement for "Secondhand Cannabis Smoke Exposure: Prevalence, Personal Use, and Neurocognitive Trajectories Over Time in Adolescents in the United States"

**Supplemental Materials**

p. 2 **Table S1.** Primary Neurocognitive Models Full Output—Pattern Comparison Processing Speed Task

p. 3 **Table S2**. Primary Neurocognitive Models Full Output— Picture Sequence Memory

Task

p. 4 **Table S3.** Primary Neurocognitive Models Full Output— Picture Vocabulary

Task

p. 5 **Table S4**. Primary Neurocognitive Models Full Output— Oral Reading

Task

p. 6 **Table S5.** Model-Derived Estimates of Mean Performance on Flanker Task by SCS Group and Age

p. 7 **Table S6**. Sensitivity Analyses – Doubly robust prenatal exposure model

p. 8 **Table S7**. Secondary Neurocognitive Models Full Output— Number of Waves of Reported SCS exposure predicting Flanker Task Performance

**Table S1. Primary Neurocognitive Models Full Output—Pattern Comparison Processing Speed Task**

|  | **Pattern Comparison Processing Speed Task (NIH Toolbox): Uncorrected Standard Score** | | |
| --- | --- | --- | --- |
| *Predictors* | *Estimates* | *CI* | *p* |
| (Intercept) | 40.52 | 36.15 – 44.90 | **<0.001** |
| SCS Status | -1.35 | -13.00 – 10.29 | 0.820 |
| Visit information: Youth's age at the start of the event | 5.04 | 4.73 – 5.34 | **<0.001** |
| Participant's sex: Female | 3.21 | 1.50 – 4.93 | **<0.001** |
| Past Year Alcohol | 3.08 | -0.35 – 6.51 | 0.078 |
| Past Year Nicotine | -3.47 | -7.13 – 0.20 | 0.064 |
| Past Year Other Substance Use | -5.21 | -20.70 – 10.28 | 0.510 |
| SCS*Age | 0.03 | -0.73 – 0.80 | 0.929 |
| **Random Effects** | | | |
| σ^2^ | 125.07 | | |
| τ_00_ _participant id_ | 74.78 | | |
| τ_00_ _family id_ | 82.34 | | |
| ICC | 0.56 | | |
| N _participant id_ | 1165 | | |
| N _family id_ | 1065 | | |
| Observations | 2646 | | |
| Marginal R^2^ / Conditional R^2^ | 0.231 / 0.659 | | |

**Table S2. Primary Neurocognitive Models Full Output— Picture Sequence Memory
Task**

|  | **Picture Sequence Memory Task (NIH Toolbox): Uncorrected Standard Score** | | |
| --- | --- | --- | --- |
| *Predictors* | *Estimates* | *CI* | *p* |
| (Intercept) | 96.42 | 92.72 – 100.12 | **<0.001** |
| SCS Status | -8.57 | -18.31 – 1.16 | 0.084 |
| Visit information: Youth's age at the start of the event | 0.93 | 0.67 – 1.20 | **<0.001** |
| Participant's sex: Female | 0.97 | -0.34 – 2.27 | 0.147 |
| Past Year Alcohol | 1.30 | -1.73 – 4.33 | 0.399 |
| Past Year Nicotine | -0.45 | -3.67 – 2.77 | 0.785 |
| Past Year Other Substance Use | -1.85 | -15.88 – 12.18 | 0.796 |
| SCS*Age | 0.55 | -0.10 – 1.19 | 0.098 |
| **Random Effects** | | | |
| σ^2^ | 114.30 | | |
| τ_00_ _participant id_ | 26.70 | | |
| τ_00_ _family id_ | 60.20 | | |
| ICC | 0.43 | | |
| N _participant id_ | 1219 | | |
| N _family id_ | 1117 | | |
| Observations | 3166 | | |
| Marginal R^2^ / Conditional R^2^ | 0.018 / 0.442 | | |

**Table S3. Primary Neurocognitive Models Full Output— Picture Vocabulary
Task**

|  | **Picture Vocabulary Task (NIH Toolbox): Uncorrected Standard Score** | | |
| --- | --- | --- | --- |
| *Predictors* | *Estimates* | *CI* | *p* |
| (Intercept) | 64.70 | 63.08 – 66.32 | **<0.001** |
| SCS Status | -3.78 | -7.97 – 0.40 | 0.076 |
| Visit information: Youth's age at the start of the event | 2.01 | 1.90 – 2.12 | **<0.001** |
| Participant's sex: Female | -0.01 | -0.86 – 0.85 | 0.987 |
| Past Year Alcohol | 0.33 | -0.96 – 1.62 | 0.618 |
| Past Year Nicotine | -0.64 | -2.03 – 0.75 | 0.367 |
| Past Year Other Substance Use | 7.52 | 1.50 – 13.54 | **0.014** |
| SCS*Age | 0.20 | -0.07 – 0.48 | 0.150 |
| **Random Effects** | | | |
| σ^2^ | 18.64 | | |
| τ_00_ _participant id_ | 8.11 | | |
| τ_00_ _family id_ | 45.04 | | |
| ICC | 0.74 | | |
| N _participant id_ | 1220 | | |
| N _family id_ | 1118 | | |
| Observations | 3172 | | |
| Marginal R^2^ / Conditional R^2^ | 0.151 / 0.780 | | |

**Table S4. Primary Neurocognitive Models Full Output— Oral Reading
Task**

|  | **Oral Reading Recognition Task (NIH Toolbox): Uncorrected Standard Score** | | |
| --- | --- | --- | --- |
| *Predictors* | *Estimates* | *CI* | *p* |
| (Intercept) | 74.58 | 73.24 – 75.92 | **<0.001** |
| SCS Status | -2.48 | -5.97 – 1.02 | 0.165 |
| Visit information: Youth's age at the start of the event | 1.68 | 1.58 – 1.77 | **<0.001** |
| Participant's sex: Female | 0.08 | -0.63 – 0.80 | 0.816 |
| Past Year Alcohol | 0.03 | -1.04 – 1.11 | 0.951 |
| Past Year Nicotine | -0.41 | -1.56 – 0.74 | 0.487 |
| Past Year Other Substance Use | -4.45 | -9.43 – 0.53 | 0.080 |
| SCS*Age | 0.14 | -0.09 – 0.37 | 0.224 |
| **Random Effects** | | | |
| σ^2^ | 12.78 | | |
| τ_00_ _participant id_ | 11.46 | | |
| τ_00_ _family id_ | 23.46 | | |
| ICC | 0.73 | | |
| N _participant id_ | 1219 | | |
| N _family id_ | 1117 | | |
| Observations | 3164 | | |
| Marginal R^2^ / Conditional R^2^ | 0.157 / 0.774 | | |

**Table S5. Model-Derived Estimates of Mean Performance on Flanker Task by SCS Group and Age**

| **Flanker Inhibitory Control and Attention Task** | **Controls** | **SCS Group** |
| --- | --- | --- |
| Age 10 | 98.33 (6.34) | 98.00 (4.94) |
| Age 11 | 99.20 (8.05) | 100.77 (6.09) |
| Age 12 | 100.30 (7.40) | 100.45 (8.04) |
| Age 13 | 102.66 (8.67) | 102.46 (8.61) |
| Age 14 | 104.15 (8.31) | 103.58 (8.39) |
| Age 15 | 105.50 (7.54) | 104.81 (7.72) |
| Age 16 | 107.00 (7.17) | 105.90 (7.31) |
| Age 17 | 108.59 (7.46) | 108.00 (5.11) |

**Table S6. Sensitivity Analyses – Doubly robust prenatal exposure model**

Given imperfect matching on prenatal cannabis exposure (SMD > .1), a sensitivity analysis was including adding prenatal cannabis exposure as a covariate to confirm results persisted.

|  | **Flanker Inhibitory Control and Attention Task (NIH Toolbox): Uncorrected Standard Score** | | |
| --- | --- | --- | --- |
| *Predictors* | *Estimates* | *CI* | *p* |
| (Intercept) | 81.03 | 78.85 – 83.21 | **<0.001** |
| SCS Status | 6.48 | 0.77 – 12.18 | **0.026** |
| Visit information: Youth's age at the start of the event | 1.61 | 1.46 – 1.77 | **<0.001** |
| Participant's sex: Female | -0.87 | -1.64 – -0.10 | **0.027** |
| Past Year Alcohol | 1.04 | -0.64 – 2.72 | 0.224 |
| Past Year Nicotine | -0.19 | -2.01 – 1.62 | 0.836 |
| Past Year Other Substance Use | -0.27 | -7.95 – 7.41 | 0.945 |
| Prenatal Cannabis Exposure | -1.38 | -2.35 – -0.41 | **0.006** |
| SCS*Age | -0.45 | -0.83 – -0.07 | **0.019** |
| **Random Effects** | | | |
| σ^2^ | 32.20 | | |
| τ_00_ _participant id_ | 18.22 | | |
| τ_00_ _family id_ | 10.37 | | |
| ICC | 0.47 | | |
| N _participant id_ | 1165 | | |
| N _family id_ | 1065 | | |
| Observations | 2656 | | |
| Marginal R^2^ / Conditional R^2^ | 0.121 / 0.535 | | |

**Table S7. Secondary Neurocognitive Models Full Output— Number of Waves of Reported SCS exposure predicting Flanker Task Performance**

|  | **Flanker Inhibitory Control and Attention Task (NIH Toolbox): Uncorrected Standard Score** | | |
| --- | --- | --- | --- |
| *Predictors* | *Estimates* | *CI* | *p* |
| (Intercept) | 81.17 | 79.08 – 83.25 | **<0.001** |
| Number of Reported SCS Waves | 3.86 | -0.31 – 8.02 | 0.069 |
| Visit information: Youth's age at the start of the event | 1.58 | 1.44 – 1.73 | **<0.001** |
| Participant's sex: Female | -0.89 | -1.66 – -0.11 | **0.025** |
| Past Year Alcohol | 1.07 | -0.61 – 2.75 | 0.210 |
| Past Year Nicotine | -0.17 | -1.99 – 1.65 | 0.855 |
| Past Year Other Substance Use | -0.10 | -7.79 – 7.58 | 0.979 |
| SCS Waves*Age | -0.25 | -0.52 – 0.01 | 0.057 |
| **Random Effects** | | | |
| σ^2^ | 32.18 | | |
| τ_00_ _participant id_ | 18.08 | | |
| τ_00_ _family id_ | 10.89 | | |
| ICC | 0.47 | | |
| N _participant id_ | 1165 | | |
| N _family id_ | 1065 | | |
| Observations | 2656 | | |
| Marginal R^2^ / Conditional R^2^ | 0.116 / 0.535 | | |
